# The accuracy of anal self- and companion exams among sexual minority men and transgender women: The Prevent Anal Cancer Palpation Study

**DOI:** 10.1101/2023.10.19.23297209

**Authors:** Alan G. Nyitray, Timothy L. McAuliffe, Cameron Liebert, Michael D. Swartz, Ashish A. Deshmukh, Elizabeth Y. Chiao, Lou Weaver, Ellen Almirol, Jared Kerman, John A. Schneider, J. Michael Wilkerson, Lu-Yu Hwang, Derek Smith, Aniruddha Hazra, The Prevent Anal Cancer Palpation Study Team

**Author notes:** **Corresponding author:** Dr. Alan G. Nyitray Clinical Cancer Center and Center for AIDS Intervention Research Medical College of Wisconsin 2071 North Summit Avenue Milwaukee, WI 53202.

## Abstract

**Background:** Squamous cell carcinoma of the anus (SCCA) annual incidence among sexual minority men (SMM) with and without HIV is 85/100,000 and 19/100,000 persons, respectively, which is significantly higher than the overall incidence (2/100,000). Since SCCA tumours average ≥30 mm at diagnosis, we assessed the accuracy of individuals to self-detect anal abnormalities.

**Methods:** The study enrolled 714 SMM and transgender women (SMM/TW), aged 25 to 81 years, in Chicago, Illinois and Houston, Texas during 2020-2022. Individuals were taught the anal self- and companion examinations (ASE/ACE). Then, a clinician performed a digital anal rectal examination (DARE) before participants conducted the ASE or ACE. Accuracy was measured along with factors associated with ASE/ACE and DARE concordance.

**Findings:** The median age was 40 years (interquartile range, 32-54), 36.8% were living with HIV, and 47.0%, 23.4%, and 23.0% were non-Hispanic white, non-Hispanic Black, and Hispanic. Clinicians detected 245 individuals with abnormalities (median diameter 3 mm). Sensitivity and specificity of the ASE/ACE was 59.6% (95%CI 53.5-65.7%) and 80.2% (95%CI 76.6-83.8%), respectively. Overall concordance was 0.73 (95% CI 0.70-0.76) between ASE/ACE and DARE and increased with increasing anal canal lesion size (p=0.02). However, concordance was lower for participants aged ≥55 years (compared to 25-34 years) and when the ASE/ACE trainer was a lay person rather than a clinician.

**Interpretation:** SMM/TW who complete an ASE or ACE are likely to detect SCCA at an early stage when malignant lesions are much smaller than the current median dimension at presentation of ≥30 mm.

**Funding:** National Cancer Institute

**Research in context:** *Evidence before this study:* While squamous cell carcinoma of the anus (SCCA) incidence is substantially elevated in people with HIV, there are currently no consensus recommendations on how to screen for it, nor is there widespread technological infrastructure for one prevailing method, high-resolution anoscopy. In the absence of screening programs, the size of SCCA tumours at diagnosis are > 30 mm. We searched PubMed for articles between January 1, 2000 and June 15, 2023 using the search terms ‘anus neoplasm’ and ‘self-examination’. We found no studies assessing the accuracy of self-examinations to detect anal masses other than our prior feasibility study.

*Added value of this study:* The primary goal of the Prevent Anal Cancer Palpation Study was to assess the accuracy of lay self-examinations and companion examinations to recognise abnormalities in the anal region. Clinicians conducted a digital anal rectal examination and recorded all lesions observed at the perianus or anal canal. The median size of lesions was 3 mm. Participants conducted lay examinations and these results were judged against a clinician’s examination. The sensitivity and specificity of the lay examinations, for any lesion at the anal canal or perianal region was 59.6% and 80.1%, respectively. As lesions increased in size, concordance increased between clinician’s exam and the lay exam.

*Implications of all the available evidence:* It is now known that high-resolution anoscopy can reduce the risk for SCCA but the infrastructure using this technology is very limited in high-resource settings and almost non-existent in low resource settings, especially where HIV prevalence is highest. The evidence suggests that self- and partner examination of the anal region is feasible and that lay persons can detect lesions that are much smaller than the prevailing size of SCCA tumours.

## INTRODUCTION

Squamous cell carcinoma of the anus (SCCA) has been recognized as one of the fastest accelerating causes of cancer incidence and death rates in the United States, with an annual increase of nearly 3%/year.^1^ This disease disproportionately affects people with HIV, especially sexual minority men (SMM) among whom annual incidence is 85/100,000 persons. Incidence is also increased among HIV-negative SMM (19/100,000 persons), and HIV-negative SMM aged ≥60 years (34/100,000).^2,3^

While there are no consensus screening recommendations for SCCA, the United States Preventive Services Task Force is currently assessing the potential role of digital anal rectal examinations (DARE), anal cytology, HPV DNA screening, and high-resolution anoscopy (HRA).^4^ An annual DARE is currently recommended by the Centers for Disease Control and Prevention^5^ and several other organizations,^6,7^ in part, because most anal cancers present with a palpable or visible tumour.^8^ These lesions are often large in retrospective studies, with a mean size of 32 mm in a 2023 Australian study^9^ and a median of 30 mm in a 2020 Chinese study.^10^ Tumour size is a strong predictor of survival and recurrence.^11^ At diagnosis, 45.5% and 44.8% of cases are at a localized or regional/distant stage.^12^

Due to SCCA tumour size and palpability, it seems quite possible that individuals could self-detect tumours through palpation of visualization of lesions.^13^ If so, self-detection could downstage SCCA, especially if it is an incentive to seek care for those who may be less likely to screen for precancers, e.g., due to poor HRA infrastructure or embarrassment.^1,14^ Previously, we established the feasibility of teaching anal self-examinations and anal companion examinations (ASE/ACE) in addition to their preliminary accuracy estimates.^15^ In the current study, we report the results of the primary objective which was to compare individuals’ ASE or ACE result with a clinician gold-standard DARE at the baseline visit to establish sensitivity and specificity for these exams and to determine factors associated with concordance between lay and clinician exams.

## METHODS

### Recruitment and protocol

The Prevent Anal Cancer (PAC) Palpation Study (NCT04090060) is assessing the accuracy of anal self-examinations and anal companion examinations based in Chicago, Illinois and Houston, Texas, USA. The study used a defined protocol in each city with study procedures approved by human protections committees at each participating institution.

Study recruitment occurred primarily through social media (i.e., Scruff, Growlr, Facebook, and Instagram), friends, in-clinic advertising, and flyers placed in gay businesses from January 2020 to December 2022. Individuals were included if they were cisgender or transgender sexual minority men or transgender women (SMM/TW), aged >25 years, and reported sex with men (cis or trans) in the last five years (or identified with a minority sexual orientation). Individuals were excluded if they had an unresolved medical diagnosis of anal condyloma, haemorrhoids, or SCCA or if they reported a DARE in the prior 3 months. People speaking English were included in Chicago, while both English and Spanish speakers were included in Houston. Recruitment was stratified to enrol single individuals and partners in couples.

Interested individuals could use a URL or QR code to access the study website which included a link for the eligibility survey. Prior to the COVID-19 pandemic eligible individuals attended a face-to-face consenting session followed by ASE/ACE instruction and assessment of exam accuracy. The study was suspended due to COVID-19 pandemic stay-at-home orders on March 14, 2020. When the study was resumed on July 30, 2020, in Chicago and October 1, 2020, in Houston, individuals were consented virtually,^16^ were asked if they wanted to participate as an individual or a couple, and then were scheduled for an appointment for ASE/ACE instruction and assessment of exam accuracy.

Almost all study appointments in Chicago occurred at a community centre for Black SMM/TW (97.0%). In Houston the appointment occurred at private clinic serving LGBTQ communities (80.3%) and a public HIV clinic (19.7%). In the original protocol all appointments were to occur at the Houston public HIV clinic; however, the COVID-19 pandemic required having most visits at the private LGBTQ clinic. The pandemic also affected recruitment: In Chicago, a majority was recruited through social media (64.1%) while in Houston, a plurality was recruited through clinics (42.8%) (Supplemental Table 1).

Physicians and advanced practice providers (APP) were trained in DARE technique and recording of findings. They practiced on mannequins and were observed by senior physicians (JAS and EYC) when conducting the initial DAREs on participants. In Chicago, a physician (AH) conducted DARE with 81.4% of participants with the remainder conducted by an APP. In Houston, only APPs conducted DARE with 72.9% of DAREs conducted by one APP (DS). Study staff tested HCPs’ assessment of lesion size on occasion by having the HCP palpate a latex lesion and guess its size without looking at the lesion.

The ASE/ACE training for study participants consisted of education about anal anatomy, anal cancer, and conducting an ASE/ACE. Trainers were usually non-clinicians (88.0%), for example, project coordinators. Accompanying illustrated instructions taught participants to use a mirror or take a selfie to view the perianus (for ASE participants only), and to palpate the entire 360° of the anal canal. Participants were taught to insert their index finger as deep as the second knuckle (proximal interphalangeal joint) since the full length of the anal canal (3-5 cm) is shorter than the male index finger.^17^ Individuals practiced the exam on two mannequins (Kyoto Kagaku Co., Kyoto, Japan), one with no anal canal lesion, and the other a 7 mm tumour. The training emphasized that the goal was only to detect if an abnormality was present (yes or no), regardless of type (e.g., wart, haemorrhoid, fissure, or tumour); thus, we assessed ASE and ACE for multiple anal conditions with the logic that if a mass or induration, regardless of aetiology, can be palpated or seen, then malignant tumours may be recognized too. Participants were told that if they found an abnormality, it was very unlikely to be cancer because other conditions like haemorrhoids and condyloma were much more common. Prostate and distal rectal palpation were not taught.^18^ The training lasted an average of 14 minutes.

The HCP then collected an anal swab and performed a DARE according to published guidelines.^19^ The HCP used the DARE as a teachable moment, e.g., “I’m feeling all 360° of your anal canal.” The HCP withheld providing DARE results until later in the visit. In addition to recording presence of an abnormality, HCPs also recorded location, size, and other characteristics of up to four abnormalities per person.

Participants were left alone in the exam room to complete the ASE or ACE and record presence of an abnormality. Participants completed the exam in an average of 4 minutes. Finally, the individual or couple completed a post-exam survey, received their DARE results and any needed clinical referrals, and were paid $50.

### Statistical methods

The central hypothesis was that the ASE/ACE would have ≥70% sensitivity and ≥90% specificity using the clinician DARE as the gold standard. The sensitivity of DARE has been estimated at 90%.^20^ Sensitivity of the ASE/ACE was stratified by the following variables and 95% confidence intervals (CI) examined by strata for overlap: type of exam (ASE vs ACE), anatomic site (anal canal vs perianal), HIV, waist size, obesity, lesions needing referral (vs. those needing no follow up), and self-reported dexterity-related comorbidities (i.e., arthritis, carpal tunnel syndrome, cerebral palsy, diabetes, fibromyalgia, chronic lower back pain, motor neuron diseases, movement disorders, multiple sclerosis, obesity, spina bifida, spinal cord injury, or stroke). All hypothesis tests were two-sided with a 0.05 alpha. We estimated concordance, positive predictive value, and negative predictive value. For couples, each partner performed digital insertion on the other partner; thus, couples provided two participant results while the clinician produced two associated DARE results.

Factors associated with participant and HCP exam agreement, i.e., concordance, were assessed. For individuals, concordance was coded as one if the result of both HCP and participant was the same and zero if clinician and participant disagreed. For partners, concordance was coded as one if both clinician and the partner performing digital insertion agreed on the results for the other partner and zero if the performing partner and clinician disagreed. For analysis of factors associated with concordance, prevalence ratios (PR), adjusted PR (aPR), and 95% CIs were calculated by bivariate and multivariable Poisson regression using a robust sandwich estimator of the variance.^21^ Variables were included in multivariable analysis if they were significant in bivariate analysis at p<0.15. Age and city were included in all modelling as a potential confounder. Waist circumference was categorized as ≤102 cm and >102 cm in accordance with prior literature identifying increased health-related problems among individuals with >102 cm waist size.^22^ Bivariate and multivariable analyses were also conducted for only those engaging in ASE (n=658).

Given a null value for sensitivity and specificity of 50% (since we sought to rule out accuracy due only to chance), our a priori power assessment indicated that power to detect sensitivity of ≥70% among 600 individuals was 0.90 and 0.81 among 100 couples using α=0.05 and two-sided tests. Power to detect specificity ≥90% among both individuals and couples was >0.90. We assumed that 11% of participants would have an abnormality.

Analyses were conducted using SAS 9.4 TS Level 1 M6 (SAS Institute, Cary, NC).

## RESULTS

Of 954 individuals consenting virtually, 236 (24.7%) did not attend the baseline study visit (Supplemental Figure). Individuals recruited through social media were less likely to attend the baseline visit compared to those recruited through advertisements or clinics (p=0.001). No other variables were associated with non-attendance at baseline. Of those who attended visit 1, the HCP was unable to complete the DARE for one person, and three individuals did not acknowledge completing the exam leaving 714 participants for analysis.

A total of 658 participants completed an ASE and 28 couples (56 partners) completed an ACE (Table 1). The median age was 40 years (range, 25-81), and 47.0%, 23.4%, and 23.0% identified as non-Hispanic white, non-Hispanic Black, and Hispanic, respectively. A total of 36.8% self-reported as a person with HIV (PWH).

**Table 1.**
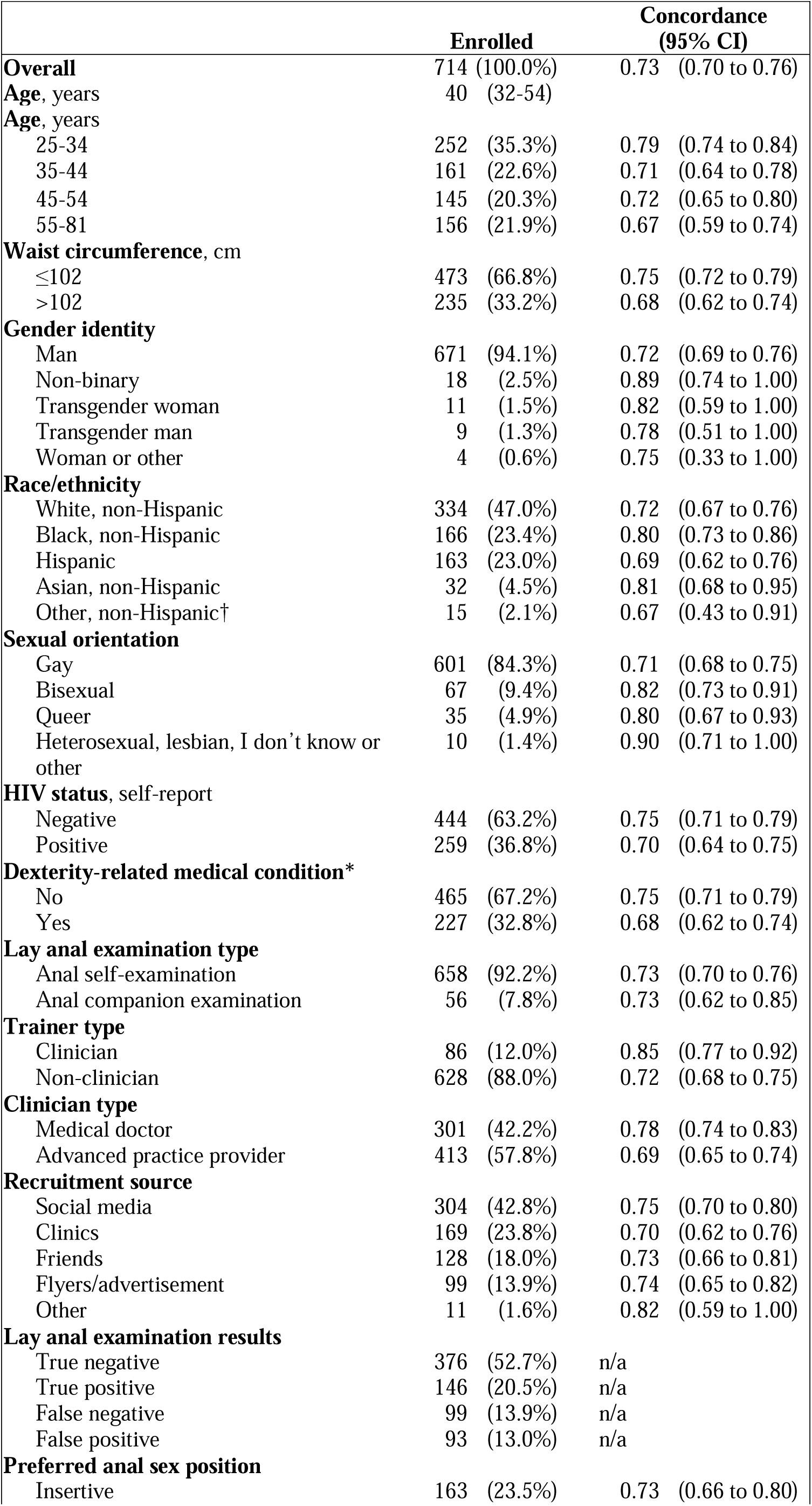
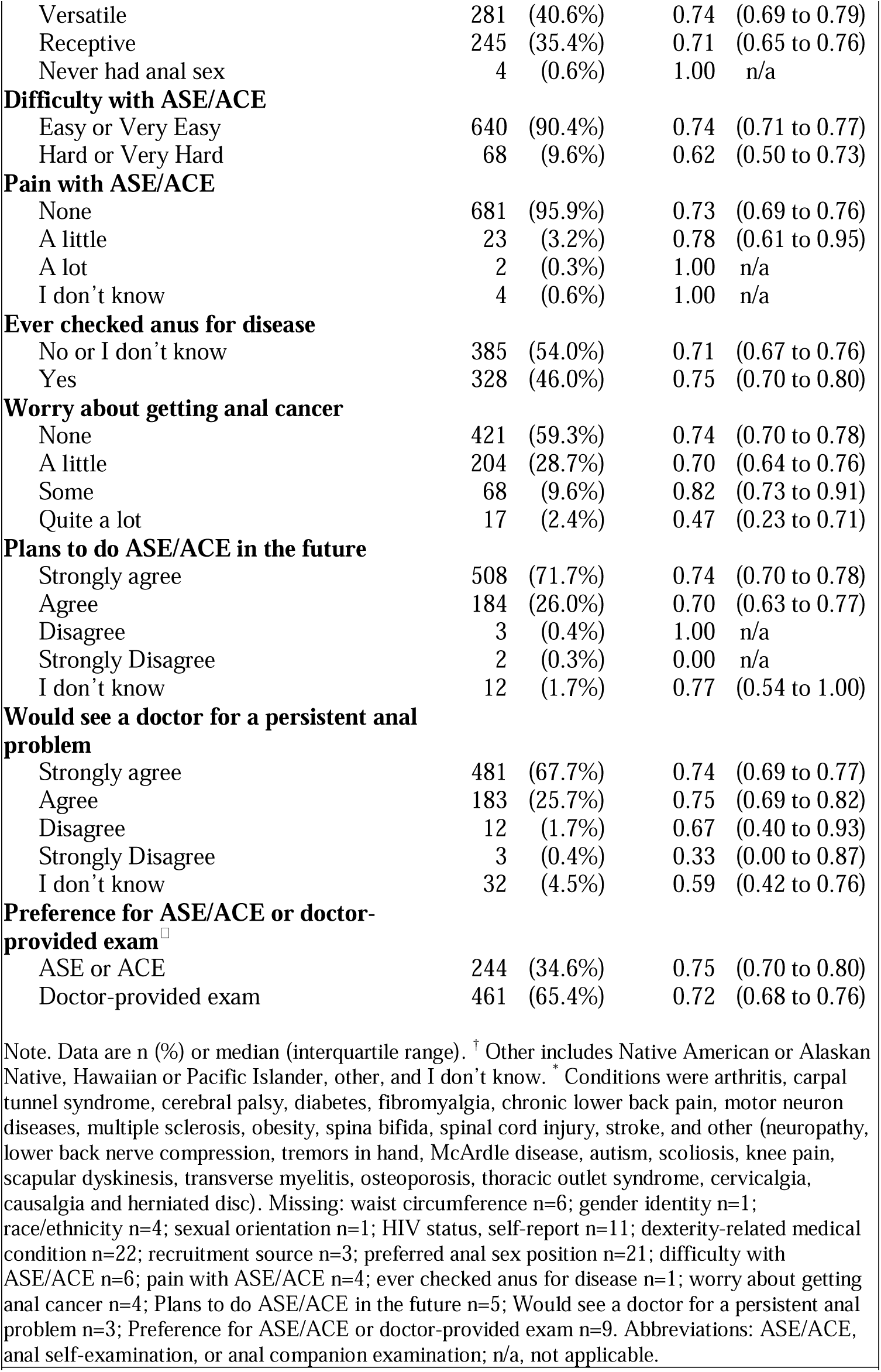
Characteristics of individuals conducting lay anal examinations and concordance with clinician examinations in Chicago, Illinois and Houston, Texas, 2020-2022.

|  | Enrolled | Concordance<br>(95% CI) |  |
| --- | --- | --- | --- |
| **Overall** | 714 (100.0%) | 0.73 | (0.70 to 0.76) |
| **Age, years** | 40 (32-54) |  |  |
| **Age, years** |  |  |  |
| 25-34 | 252 (35.3%) | 0.79 | (0.74 to 0.84) |
| 35-44 | 161 (22.6%) | 0.71 | (0.64 to 0.78) |
| 45-54 | 145 (20.3%) | 0.72 | (0.65 to 0.80) |
| 55-81 | 156 (21.9%) | 0.67 | (0.59 to 0.74) |
| **Waist circumference, cm** |  |  |  |
| ≤102 | 473 (66.8%) | 0.75 | (0.72 to 0.79) |
| >102 | 235 (33.2%) | 0.68 | (0.62 to 0.74) |
| **Gender identity** |  |  |  |
| Man | 671 (94.1%) | 0.72 | (0.69 to 0.76) |
| Non-binary | 18 (2.5%) | 0.89 | (0.74 to 1.00) |
| Transgender woman | 11 (1.5%) | 0.82 | (0.59 to 1.00) |
| Transgender man | 9 (1.3%) | 0.78 | (0.51 to 1.00) |
| Woman or other | 4 (0.6%) | 0.75 | (0.33 to 1.00) |
| **Race/ethnicity** |  |  |  |
| White, non-Hispanic | 334 (47.0%) | 0.72 | (0.67 to 0.76) |
| Black, non-Hispanic | 166 (23.4%) | 0.80 | (0.73 to 0.86) |
| Hispanic | 163 (23.0%) | 0.69 | (0.62 to 0.76) |
| Asian, non-Hispanic | 32 (4.5%) | 0.81 | (0.68 to 0.95) |
| Other, non-Hispanic† | 15 (2.1%) | 0.67 | (0.43 to 0.91) |
| **Sexual orientation** |  |  |  |
| Gay | 601 (84.3%) | 0.71 | (0.68 to 0.75) |
| Bisexual | 67 (9.4%) | 0.82 | (0.73 to 0.91) |
| Queer | 35 (4.9%) | 0.80 | (0.67 to 0.93) |
| Heterosexual, lesbian, I don't know or other | 10 (1.4%) | 0.90 | (0.71 to 1.00) |
| **HIV status, self-report** |  |  |  |
| Negative | 444 (63.2%) | 0.75 | (0.71 to 0.79) |
| Positive | 259 (36.8%) | 0.70 | (0.64 to 0.75) |
| **Dexterity-related medical condition\*** |  |  |  |
| No | 465 (67.2%) | 0.75 | (0.71 to 0.79) |
| Yes | 227 (32.8%) | 0.68 | (0.62 to 0.74) |
| **Lay anal examination type** |  |  |  |
| Anal self-examination | 658 (92.2%) | 0.73 | (0.70 to 0.76) |
| Anal companion examination | 56 (7.8%) | 0.73 | (0.62 to 0.85) |
| **Trainer type** |  |  |  |
| Clinician | 86 (12.0%) | 0.85 | (0.77 to 0.92) |
| Non-clinician | 628 (88.0%) | 0.72 | (0.68 to 0.75) |
| **Clinician type** |  |  |  |
| Medical doctor | 301 (42.2%) | 0.78 | (0.74 to 0.83) |
| Advanced practice provider | 413 (57.8%) | 0.69 | (0.65 to 0.74) |
| **Recruitment source** |  |  |  |
| Social media | 304 (42.8%) | 0.75 | (0.70 to 0.80) |
| Clinics | 169 (23.8%) | 0.70 | (0.62 to 0.76) |
| Friends | 128 (18.0%) | 0.73 | (0.66 to 0.81) |
| Flyers/advertisement | 99 (13.9%) | 0.74 | (0.65 to 0.82) |
| Other | 11 (1.6%) | 0.82 | (0.59 to 1.00) |
| **Lay anal examination results** |  |  |  |
| True negative | 376 (52.7%) | n/a |  |
| True positive | 146 (20.5%) | n/a |  |
| False negative | 99 (13.9%) | n/a |  |
| False positive | 93 (13.0%) | n/a |  |
| **Preferred anal sex position** |  |  |  |
| Insertive | 163 (23.5%) | 0.73 | (0.66 to 0.80) |

|  |  |  |
| --- | --- | --- |
| Versatile | 281 (40.6%) | 0.74 (0.69 to 0.79) |
| Receptive | 245 (35.4%) | 0.71 (0.65 to 0.76) |
| Never had anal sex | 4 (0.6%) | 1.00 n/a |
| **Difficulty with ASE/ACE** |  |  |
| Easy or Very Easy | 640 (90.4%) | 0.74 (0.71 to 0.77) |
| Hard or Very Hard | 68 (9.6%) | 0.62 (0.50 to 0.73) |
| **Pain with ASE/ACE** |  |  |
| None | 681 (95.9%) | 0.73 (0.69 to 0.76) |
| A little | 23 (3.2%) | 0.78 (0.61 to 0.95) |
| A lot | 2 (0.3%) | 1.00 n/a |
| I don't know | 4 (0.6%) | 1.00 n/a |
| **Ever checked anus for disease** |  |  |
| No or I don't know | 385 (54.0%) | 0.71 (0.67 to 0.76) |
| Yes | 328 (46.0%) | 0.75 (0.70 to 0.80) |
| **Worry about getting anal cancer** |  |  |
| None | 421 (59.3%) | 0.74 (0.70 to 0.78) |
| A little | 204 (28.7%) | 0.70 (0.64 to 0.76) |
| Some | 68 (9.6%) | 0.82 (0.73 to 0.91) |
| Quite a lot | 17 (2.4%) | 0.47 (0.23 to 0.71) |
| **Plans to do ASE/ACE in the future** |  |  |
| Strongly agree | 508 (71.7%) | 0.74 (0.70 to 0.78) |
| Agree | 184 (26.0%) | 0.70 (0.63 to 0.77) |
| Disagree | 3 (0.4%) | 1.00 n/a |
| Strongly Disagree | 2 (0.3%) | 0.00 n/a |
| I don't know | 12 (1.7%) | 0.77 (0.54 to 1.00) |
| **Would see a doctor for a persistent anal problem** |  |  |
| Strongly agree | 481 (67.7%) | 0.74 (0.69 to 0.77) |
| Agree | 183 (25.7%) | 0.75 (0.69 to 0.82) |
| Disagree | 12 (1.7%) | 0.67 (0.40 to 0.93) |
| Strongly Disagree | 3 (0.4%) | 0.33 (0.00 to 0.87) |
| I don't know | 32 (4.5%) | 0.59 (0.42 to 0.76) |
| **Preference for ASE/ACE or doctor-provided exam<sup>†</sup>** |  |  |
| ASE or ACE | 244 (34.6%) | 0.75 (0.70 to 0.80) |
| Doctor-provided exam | 461 (65.4%) | 0.72 (0.68 to 0.76) |

Note. Data are n (%) or median (interquartile range). <sup>†</sup> Other includes Native American or Alaskan Native, Hawaiian or Pacific Islander, other, and I don't know. \* Conditions were arthritis, carpal tunnel syndrome, cerebral palsy, diabetes, fibromyalgia, chronic lower back pain, motor neuron diseases, multiple sclerosis, obesity, spina bifida, spinal cord injury, stroke, and other (neuropathy, lower back nerve compression, tremors in hand, McArdle disease, autism, scoliosis, knee pain, scapular dyskinesis, transverse myelitis, osteoporosis, thoracic outlet syndrome, cervicalgia, causalgia and herniated disc). Missing: waist circumference n=6; gender identity n=1; race/ethnicity n=4; sexual orientation n=1; HIV status, self-report n=11; dexterity-related medical condition n=22; recruitment source n=3; preferred anal sex position n=21; difficulty with ASE/ACE n=6; pain with ASE/ACE n=4; ever checked anus for disease n=1; worry about getting anal cancer n=4; Plans to do ASE/ACE in the future n=5; Would see a doctor for a persistent anal problem n=3; Preference for ASE/ACE or doctor-provided exam n=9. Abbreviations: ASE/ACE, anal self-examination, or anal companion examination; n/a, not applicable.

HCPs recorded ≥1 abnormality in 245 individuals (34.3%) with a median diameter at both anal canal and perianal region of 3 mm for the primary lesion (anal canal lesion size range, 1 – 8 mm; perianal lesion size range, 1 – 10 mm) (Table 2). The most prevalent lesion types appearing in the anal canal and perianus were described by HCPs as haemorrhoids (46.8%) and skin folds, flaps, or tags (47.4%), respectively.

**Table 2.** Lesion characteristics for 245 individuals in Chicago, Illinois and Houston, Texas, 2020-2022.

|  | **Perianus (n=190)\*** | **Anal canal (n=94)\*** |
| --- | --- | --- |
| **Lesion size, mm** | 3, 1-10 | 3, 1-8 |
| **Lesion type** |  |  |
| Haemorrhoids | 53 (27.9%) | 44 (46.8%) |
| Skin fold/flaps/tags | 90 (47.4%) | 6 (6.4%) |
| Scar | 12 (6.3%) | 14 (14.9%) |
| Condyloma | 14 (7.4%) | 9 (9.6%) |
| Suspicious lump or thickening | 2 (1.1%) | 14 (14.9%) |
| Papule | 9 (4.7%) | 6 (6.4%) |
| Anal fissure | 2 (1.1%) | 1 (1.1%) |
| Other | 5 (2.6%) | - |
| Perianal lichenification | 1 (0.5%) | - |
| Perianal psoriasis | 1 (0.5%) | - |
| Cyst | 1 (0.5%) | - |
| **HCP referred participant** |  |  |
| No | 166 (87.4%) | 64 (68.1%) |
| Yes | 24 (12.6%) | 30 (31.9%) |

Note. Data are median, range and n and (%). \* Total number of individuals with primary lesions at either site is > 245 due to lesions at both anatomic sites in some individuals. Abbreviations: HCP, health care provider.

Overall sensitivity and specificity of the ASE/ACE was 59.6% and 80.2%, respectively, while positive predictive value and negative predictive value was 61.1% and 79.2%, respectively (Figure 1). When stratified by type of exam (ASE vs ACE), anatomic site (anal canal vs perianus), waist size, lesions needing referral (vs. those needing no follow up), and self-reported dexterity-related comorbidities, no significant differences in sensitivity were observed. If a lesion prompted a clinician’s referral, sensitivity was 63.0% (Supplementary Table 1).

**Figure 1.** Agreement and accuracy for lay anal examinations compared with clinician examinations in Chicago, Illinois and Houston, Texas, 2020-2022.

The ASE/ACE had 0.73 concordance with the clinician DARE. The ASE/ACE result was a true negative for 52.7% of all exams and true positive for an additional 20.5% of exams (Table 1). The ASE/ACE were discordant in 0.27 of exams (false negative 13.9%; false positive 13.0%).

Concordance for anal canal lesions increased with increasing lesion size (p=0.02, Figure 2). Concordance was not associated with other variables except for trainer type, clinician type, and amount of anal cancer worry (Table 1). Concordance was 0.85 (95% CI 0.77-0.92) when clinicians conducted the training and 0.72 (95% CI 0.68-0.75) when a non-clinician conducted the training. Concordance was 0.78 (95% CI 0.74-0.83) when a physician conducted the DARE and 0.69 when an APP conducted the DARE (95% CI 0.65-0.74). While only 17 participants reported worrying “quite a lot” about anal cancer, they had lower concordance than participants who said they worried “some” about anal cancer (0.47, 95% CI 0.23-0.71 for “quite a lot” and 0.82, 95% CI 0.73-0.91 for “some”).

**Figure 2.** Concordance by lesion size between lay and clinician anal examinations stratified by anatomic site in Chicago, Illinois and Houston, Texas, 2020-2022. P value is derived from the Cochrane-Armitage test for trend.

After performing the ASE or ACE, 97.7% of individuals agreed or strongly agreed they would do another in the future (Table 1). Most individuals (93.4%) said they agreed or strongly agreed they would see a doctor for a persistent anal abnormality. A total of 34.6% of individuals preferred the ASE/ACE exam versus a clinician’s DARE examination.

Almost all individuals (95.9%) said the lay exam was not painful. Of those reporting pain (n=25), 23 reported “a little” pain and 2 individuals reported “a lot” of pain. Neither of the latter two participants required follow up. In bivariate analysis, the oldest participants were less likely to be concordant with the clinician (PR 0.85, 95% CI 0.75-0.96 for 55-81 years old compared with 25-34 years old) as were participants with a waist size greater than 102 cm (PR 0.90, 95% CI 0.82-1.00, compared with waist sizes ≤ 102 cm) (Table 3). A more accurate ASE/ACE was associated with Black, non-Hispanic race/ethnicity (PR 1.11, 95% CI 1.00-1.23, compared with white, non-Hispanic individuals), and a non-gay sexual orientation. For example, individuals identifying as bisexual had increased concordance (PR 1.15, 95% CI 1.02-1.30, compared with gay). The ASE/ACE were less likely to be concordant with the clinician exam when a non-clinician conducted the training versus when a clinician conducted the training (PR 0.84, 95% CI 0.76-0.93). The ASE/ACE were also less likely to be concordant with the clinician exam when it was conducted by an APP rather than a doctor (PR 0.88, 95% CI 0.81-0.96).

**Table 3.** Factors associated with concordance between lay and clinician anal examinations in Chicago, Illinois and Houston, Texas, 2020-2022, bivariate and multivariable analyses.

| Characteristic | PR (95% CI) | aPR* (95% CI) |
| --- | --- | --- |
| **Age, years** |  |  |
| 25-34 | 1.0 | 1.0 |
| 35-44 | 0.91 (0.81-1.02) | 0.91 (0.81-1.03) |
| 45-54 | 0.92 (0.82-1.04) | 0.94 (0.84-1.07) |
| 55-81 | **0.85 (0.75-0.96)** | **0.87 (0.76-0.99)** |
| **Waist, cm** |  |  |
| ≤ 102 | 1.0 |  |
| > 102 | **0.90 (0.82-1.00)** | - |
| **Race/ethnicity** |  |  |
| White, non-Hispanic | 1.0 |  |
| Black, non-Hispanic | **1.11 (1.00-1.23)** | - |
| Hispanic | 0.96 (0.85-1.09) | - |
| Asian, non-Hispanic | 1.14 (0.95-1.36) | - |
| Other, non-Hispanic† | 0.93 (0.65-1.34) | - |
| **Sexual orientation** |  |  |
| Gay | 1.0 |  |
| Bisexual | **1.15 (1.02-1.30)** | - |
| Queer | 1.12 (0.94-1.33) | - |
| Heterosexual, lesbian, don't know or other | **1.26 (1.02-1.56)** | - |
| **HIV status, self-report** |  |  |
| Negative | 1.0 | - |
| Positive | 0.92 (0.84-1.02) | - |
| **Dexterity-related medical condition** |  |  |
| No | 1.0 | - |
| Yes | 0.91 (0.82-1.01) | - |
| **Difficulty with ASE/ACE** |  |  |
| Hard or Very Hard | 1.0 |  |
| Easy or Very Easy | 1.20 (0.99-1.45) | - |
| **Trainer type** |  |  |
| Clinician | 1.0 | 1.0 |
| Non-clinician | **0.84 (0.76-0.93)** | **0.87 (0.78-0.98)** |
| **Clinician type** |  |  |
| Medical doctor | 1.0 |  |
| Advanced practice provider | **0.88 (0.81-0.96)** | - |
| **Worry about getting anal cancer** |  |  |
| None | 1.0 |  |
| A little | 0.95 (0.85-1.06) | - |
| Some | 1.11 (0.99-1.26) | - |
| Quite a lot | 0.64 (0.38-1.06) | - |

Note. Confidence intervals in bold do not include unity. \* Variables remaining in model are adjusted for each other and city. † Other includes American Indian or Alaskan Native, Native Hawaiian or Pacific Islander, and other. Abbreviations: PR, prevalence ratio; aPR, adjusted prevalence ratio; anal self-examination, or anal companion examination.

In multivariable analysis age was inversely associated with concordance (aPR 0.87, 95% CI 0.76-0.99 for 55-81 years compared to 25-34 years). Also, when the trainer was a non-clinician there was decreased concordance with the ASE/ASE as compared to the clinician’s training (aPR 0.87, 95%CI 0.78-0.98). When the dataset was restricted to only individuals doing the ASE, the multivariable analysis yielded comparable results except that ‘some’ worry about getting anal cancer compared to no worry was associated with increased concordance in multivariable analysis (aPR 1.14, 95% CI 1.01-1.29) (Supplementary Table 3)

## DISCUSSION

Among people who are particularly vulnerable to SCCA, HCPs recorded 245 abnormalities in the anal canal and perianus with a median diameter of 3 mm and a range of 1-10 mm. After being taught how to conduct an ASE or ACE in clinics and a community drop-in centre, participants achieved 59.6% sensitivity to detect these lesions when compared with an HCP DARE. As anal canal lesion size increased there was increasing concordance between HCPs’ DARE and participants’ anal self- or companion exams. These results indicate that lay individuals who complete an ASE or ACE are likely to detect SCCA at an early stage when malignant lesions are smaller than the known median dimension at presentation of ≥ 30 mm.^10^

About one-third of individuals preferred the anal self- or companion exam rather than a DARE which may speak to embarrassment associated with DARE**^23^** and stigma regarding anal health;**^14^** thus, the ASE/ACE may increase anal cancer screening access by providing an option to detect an abnormality that motivates a clinic visit. It is worth mentioning that most HCPs are not routinely doing a DARE as recommended;**^5,24^** thus an ASE/ACE may trigger a needed clinical encounter.

To our knowledge, this study is the first to assess accuracy of anal self-examinations other than our prior feasibility study where we found higher sensitivity and specificity with 200 SMM/TW (75% and 94%, respectively).^15^ The estimate produced by the current sample of 714 SMM/TW may be closer to the ASE/ACE actual sensitivity when considered in the context of multiple clinicians and trainers, including trainers who are not clinicians. In the smaller study, one clinician provided all the education and DAREs. Since it is unlikely that clinicians have time to teach the ASE/ACE, the current study sensitivity may be closer to the truth outside of a study setting. Also, in the prior and current study, participants were more likely to palpate larger lesions. Given reports of excellent cure rates^25^ including 100% disease-specific cure rate for anal cancer tumours ≤10 mm,^26^ self-recognition of early-stage tumours (i.e., ≤20 mm) is likely to reduce anal cancer morbidity and mortality.

False-negative (13.9%) and false-positive (13.0%) results may lead to missed disease, anxiety, and unnecessary clinic visits for procedures like DARE. An annual DARE is currently recommended for PWH and HIV-negative SMM with a history of receptive anal sex. These could help address both false-negative and false-positive self- and companion exams; however, only 13.7% of PWH and HIV-negative SMM report receiving a DARE in the prior year.^24^ As with breast self-exams,^27^ it is also possible that practicing the ASE/ACE could lead to more involvement with HCPs, and thus correction of a false positive result with a clinician’s DARE.

Individual characteristics did not affect concordance except for age with individuals ≥55 years having a 13% lower likelihood of concordance compared to individuals 25-34 years. Given strong variation in anal cancer incidence by age, it is a limitation of the exams if older people at increased cancer risk are less likely to detect a lesion. Associated with biological age, we found lower concordance among individuals with larger waistlines, and dexterity-related medical conditions although the associations were not significant. We also believe it is notable that younger people had increased ability to detect lesions which may support self-detection of common anal conditions like condyloma.

Concordance also differed by trainer type with non-clinician educators having a 13% lower likelihood of concordance than clinicians. It is possible that individuals will focus more on an ASE/ACE training conducted by a clinician than a lay person. Even so, individuals’ reported self-efficacy did not differ by trainer type with >90% of participants saying that they knew how to do an ASE/ASE after being taught, that the ASE/ACE were easy to perform, that ASE/ACE can detect cancer, and that the ASE/ACE can help a person avoid cancer (data not shown).

While we exceeded our goal of enrolling participants to do the self-exam, it is a limitation that we did not reach our goal of enrolling 100 couples to do the anal companion exam. Difficulty recruiting couples was also observed in the prior study.^15^ It may be that the self-exam is preferred by most individuals rather than engaging a partner in an exam. A higher proportion of participants engaging in the ACE with a partner were more likely to be married or cohabitating than single with no partner (p<0.001) and more likely to be white, non-Hispanic compared with Black, non-Hispanic (p=0.04). Nevertheless, judged against a clinician’s exam, both ASE and ACE had the same concordance, 0.73. The companion exam may be beneficial for some people who cannot perform the ASE, for example, due to disability.

Although all clinicians received training in conducting a DARE and assessing lesion diameter, it is a limitation that clinicians did not use a measuring device and thus their assessed lesion size may not be correct. However, the primary HCP in Chicago and Houston conducted 302 and 274 DAREs, respectively, which may lead to repeatability in assessment of lesion size, but not necessarily validity.

It is now known that treatment of high-grade squamous intraepithelial (HSIL) lesions at the perianus or anal canal will reduce the risk of invasion.^28^ It seems unlikely that HSIL can be palpated although superficially invasive squamous cell carcinomas (SISCCA) of the anal canal and perianus, a minimally invasive T1 cancer with ≤ 7 mm of lateral spread, can be palpated by clinicians^8^ and thus may be palpated through self-exams too.^8,29^

Given the potential for recurrence of SCCA,^11^ ASE/ACE could also support detection of a recurring tumour in addition to submucosal tumours that evade detection with HRA. We believe it is important for individuals to establish baseline anal health with a minimum of an HCP-conducted DARE. Thereafter, repeated ASE/ACE may increase an individual’s familiarity with their anus and increase recognition of subtle changes in tissue. Finally, these exams could be used in settings that lack the option of HRA including many regions of high-resource countries and many other international contexts.^1^

Strengths of this study include the large cohort, the diversity of participants, and settings in both a clinic and a drop-in centre. In addition to previously discussed limitations, the study is also limited by COVID-19 pandemic-related protocol changes that modified consenting procedures, the clinic site in Houston, and recruitment methods.

Given high anal cancer incidence among SMM, no uniform screening standard for anal cancer, and limited infrastructure for proposed screening modalities (in both high and low-resource settings),^1^ these results suggest the ASE/ACE be further explored for detecting early-stage anal cancer when treatment is more successful.^30^ It is unknown what is the optimal frequency for performing ASE/ACE. Nevertheless, it may be a valuable community-led tool for both raising awareness and for screening for anal cancer. Given that a large proportion (46%) of SMM/TW in this study reported using their fingers in the past to check their anus for problems (without benefit of educational instruction), and given extremely high incidence of anal cancer, future research should assess the potential for implementation of ASE/ACE for populations with increased vulnerability to SCCA.

## Data Availability

De-identified data sets are available upon reasonable request to the authors.

## Author Contributions

AGN conceptualized the study and secured funding. AGN, MDS, TLM, and EYC developed the methodology. CL, LW, AH, and JK administered the clinics and taught participants how to do the lay examinations. AH and DS conducted clinical examinations. AGN, CL, and EA managed the data. AGN, and TLM conducted the formal analysis. AGN wrote the first draft of the manuscript. AGN, TLM, CL, MDS, AAD, EYC, LW, EA, JAS, JMW, LH, DS, AH, AAD reviewed and edited manuscript drafts.

## Acknowledgements

We thank the participants in the study, the Prevent Anal Cancer (PAC) Palpation Study Community Advisory Board, and the PAC Palpation Study Team: Bridgett Brzezinski, Rey Flores, DeJuan Washington, Andrew Travis, Christopher Ajala, Dr Jenna Nitkowski, Madison Humphry, Dr Gordon E. Crofoot.

## Data sharing

Fully de-identified datasets and data dictionary will be shared with properly trained investigators on the study website (https://mindyourbehind.org) within 1 year of study completion after assessment of institutional policies, Medical College of Wisconsin Human Research Protections Program rules, as well as local, state, and federal laws and regulations. Further information is available from the corresponding author upon request.

## Supplementary Material

***Supplemental Table 1.*** Stratified sensitivity estimates for lay anal examinations compared with clinician examination in Chicago, Illinois and Houston, Texas, 2020-2022

***Supplemental Table 2.*** Characteristics of individuals conducting lay anal examinations and concordance with clinician examinations by city in Chicago, Illinois and Houston, Texas, 2020-2022

***Supplemental Table 3.*** Factors associated with concordance between anal self-examinations and clinician examinations in Chicago, Illinois and Houston, Texas, 2020-2022, bivariate and multivariable analyses

***Supplemental Figure.*** Study flow

