## Supplemental Tables 1-3 and Figure for "The accuracy of anal self- and companion exams among sexual minority men and transgender women: The Prevent Anal Cancer Palpation Study"

***Supplemental Table 1.*** Stratified sensitivity estimates for lay anal examinations compared with clinician examination in Chicago, Illinois and Houston, Texas, 2020-2022

***Supplemental* *Table 2.*** Characteristics of individuals conducting lay anal examinations and concordance with clinician examinations by city in Chicago, Illinois and Houston, Texas, 2020-2022

***Supplemental Table 3.*** Factors associated with concordance between anal self-examinations and clinician examinations in Chicago, Illinois and Houston, Texas, 2020-2022, bivariate and multivariable analyses

***Supplemental* *Figure.*** Study flow

| **Stratification variable** | **Sensitivity (95% CI)** |
| --- | --- |
| **Age**, years |  |
| <45 | 64.7 (56.1-73.3) |
| ≥45 | 54.8 (46.1-63.5) |
| **Waist circumference**, cm |  |
| ≤102 | 60.4 (52.9-67.9) |
| >102 | 58.0 (47.3-68.8) |
| **HIV status**, self-report |  |
| Negative | 62.5 (54.4-70.6) |
| Positive | 56.2 (46.7-65.7) |
| **Dexterity-related medical condition** |  |
| No | 61.4 (53.7-69.2) |
| Yes | 56.5 (45.9-67.0) |
| **Preferred anal sex position** |  |
| Insertive | 52.9 (39.2-66.6) |
| Versatile | 62.4 (52.5-72.2) |
| Receptive | 60.0 (50.2-69.9) |
| Never had anal sex | 100.0 n/a |
| **Trainer type** |  |
| Clinician | 70.4 (53.2-87.6) |
| Non-Clinician | 58.3 (51.7-64.8) |
| **Lesion size** (n=245) |  |
| <5 mm | 58.4 (51.5-65.3) |
| ≥5 mm | 64.6 (51.1-78.1) |
| **Referral for any lesion** (n=245) |  |
| No | 58.8 (52.0-65.6) |
| Yes | 63.0 (49.1-77.0) |
| **Referral for anal canal lesion** (n=94) |  |
| No | 59.5 (53.0-66.1) |
| Yes | 60.0 (42.5-77.5) |
| **Referral for perianal lesion** (n=190) |  |
| No | 57.9 (51.4-64.4) |
| Yes | 75.0 (57.7-92.3) |
| Note. Numbers are percent. | |
| ***Supplemental Table 1.* Stratified sensitivity estimates for lay anal examinations compared with clinician examinations in Chicago, Illinois and Houston, Texas, 2020-2022** | |

|  | **Chicago** | **Concordance**  **(95% CI)** | **Houston** | **Concordance**  **(95% CI)** |
| --- | --- | --- | --- | --- |
| **Overall** | 370 (100.0%) | 0.76 (0.72 to 0.81) | 344 (100.0%) | 0.70 (0.65 to 0.75) |
| **Age**, years | 37 (30-51) |  | 46 (35-56) |  |
| **Age**, years |  |  |  |  |
| 25-34 | 162 (43.8%) | 0.78 (0.72 to 0.85) | 90 (26.2%) | 0.79 (0.70 to 0.87) |
| 35-44 | 92 (24.9%) | 0.79 (0.71 to 0.88) | 69 (20.1%) | 0.61 (0.49 to 0.72) |
| 45-54 | 54 (14.6%) | 0.74 (0.62 to 0.86) | 91 (26.5%) | 0.73 (0.63 to 0.82) |
| 55-81 | 62 (16.8%) | 0.68 (0.56 to 0.79) | 94 (27.3%) | 0.66 (0.56 to 0.76) |
| **Waist circumference**, cm |  |  |  |  |
| ≤102 | 262 (70.8%) | 0.81 (0.76 to 0.85) | 211 (62.4%) | 0.69 (0.63 to 0.75) |
| >102 | 108 (29.2%) | 0.66 (0.57 to 0.75) | 127 (37.6%) | 0.71 (0.63 to 0.79) |
| **Gender identity** |  |  |  |  |
| Man | 344 (93.2%) | 0.76 (0.71 to 0.80) | 327 (95.1%) | 0.69 (0.64 to 0.74) |
| Non-binary | 16 (4.3%) | 0.88 (0.71 to 1.00) | 2 (0.6%) | 1.0 n/a |
| Transgender woman | 5 (1.4%) | 0.80 (0.45 to 1.00) | 6 (1.7%) | 0.83 (0.54 to 1.00) |
| Transgender man | 1 (0.3%) | 1.0 n/a | 8 (2.3%) | 0.75 (0.45 to 1.00) |
| Woman or other | 3 (0.8%) | 0.67 (0.13 to 1.00) | 1 (0.3%) | 1.0 n/a |
| **Race/ethnicity** |  |  |  |  |
| White, non-Hispanic | 149 (40.3%) | 0.75 (0.68 to 0.82) | 185 (54.4%) | 0.70 (0.63 to 0.76) |
| Black, non-Hispanic | 120 (32.4%) | 0.79 (0.72 to 0.86) | 46 (13.5%) | 0.80 (0.69 to 0.92) |
| Hispanic | 76 (20.5%) | 0.72 (0.62 to 0.82) | 87 (25.6%) | 0.66 (0.56 to 0.76) |
| Asian, non-Hispanic | 16 (4.3%) | 0.88 (0.71 to 1.00) | 16 (4.7%) | 0.75 (0.54 to 0.96) |
| Other, non-Hispanic† | 9 (2.4%) | 0.78 (0.51 to 1.00) | 6 (1.8%) | 0.50 (0.10 to 0.90) |
| **Sexual orientation** |  |  |  |  |
| Gay | 296 (80.0%) | 0.75 (0.70 to 0.80) | 305 (88.9%) | 0.68 (0.63 to 0.73) |
| Bisexual | 40 (10.8%) | 0.83 (0.71 to 0.94) | 27 (7.9%) | 0.81 (0.67 to 0.96) |
| Queer | 28 (8.0%) | 0.75 (0.59 to 0.91) | 7 (2.0%) | 1.0 n/a |
| Heterosexual, lesbian, I don’t know or other | 6 (1.6%) | 0.83 (0.54 to 1.00) | 4 (1.2%) | 1.0 n/a |
| **HIV status**, self-report |  |  |  |  |
| Negative | 272 (65.1%) | 0.78 (0.73 to 0.83) | 172 (50.4%) | 0.72 (0.65 to 0.79) |
| Positive | 90 (24.9%) | 0.72 (0.63 to 0.81) | 169 (49.6%) | 0.68 (0.61 to 0.75) |
| **Dexterity-related medical condition*** |  |  |  |  |
| No | 255 (72.7%) | 0.77 (0.72 to 0.82) | 210 (61.6%) | 0.72 (0.66 to 0.78) |
| Yes | 96 (27.4%) | 0.72 (0.62 to 0.81) | 131 (38.4%) | 0.66 (0.58 to 0.74) |
| **Lay anal examination type** |  |  |  |  |
| Anal self-examination | 352 (95.1%) | 0.76 (0.71 to 0.80) | 306 (89.0%) | 0.70 (0.65 to 0.75) |
| Anal companion examination | 18 (4.9%) | 0.89 (0.74 to 1.00) | 38 (11.1%) | 0.68 (0.54 to 0.83) |
| **Trainer type** |  |  |  |  |
| Clinician | 84 (22.7%) | 0.86 (0.78 to 0.93) | 2 (0.6%) | 0.50 (0.00 to 1.00) |
| Non-clinician | 286 (77.3%) | 0.73 (0.68 to 0.79) | 342 (99.4%) | 0.70 (0.65 to 0.75) |
| **Clinician type** |  |  |  |  |
| Medical doctor | 301 (81.4%) | 0.78 (0.74 to 0.83) | 0 | 0.0 n/a |
| Advanced practice provider | 69 (18.7%) | 0.67 (0.56 to 0.78) | 344 (100.0%) | 0.70 (0.65 to 0.75) |
| **Recruitment source** |  |  |  |  |
| Social media | 236 (64.3%) | 0.76 (0.70 to 0.81) | 68 (19.8%) | 0.71 (0.60 to 0.81) |
| Clinics | 22 (6.0%) | 0.73 (0.54 to 0.91) | 147 (42.7%) | 0.69 (0.61 to 0.76) |
| Friends | 44 (12.0%) | 0.77 (0.65 to 0.90) | 84 (24.4%) | 0.71 (0.62 to 0.81) |
| Flyers/advertisement | 61 (16.6%) | 0.77 (0.67 to 0.88) | 38 (11.1%) | 0.68 (0.54 to 0.83) |
| Other | 4 (1.1%) | 0.75 (0.33 to 1.00) | 7 (2.0%) | 0.86 (0.60 to 1.00) |
| **Lay anal examination results** |  |  |  |  |
| True negative | 198 (53.5%) | n/a | 178 (51.7%) | n/a |
| True positive | 84 (22.7%) | n/a | 63 (18.3%) | n/a |
| False negative | 39 (10.5%) | n/a | 60 (17.4%) | n/a |
| False positive | 49 (13.2%) | n/a | 43 (12.5%) | n/a |
| **Preferred anal sex position** |  |  |  |  |
| Insertive | 91 (25.6%) | 0.77 (0.68 to 0.86) | 72 (21.4%) | 0.69 (0.59 to 0.80) |
| Versatile | 137 (38.5%) | 0.74 (0.67 to 0.82) | 144 (42.7%) | 0.73 (0.66 to 0.80) |
| Receptive | 127 (35.7%) | 0.75 (0.67 to 0.82) | 118 (35.0%) | 0.66 (0.58 to 0.75) |
| Never had anal sex | 1 (0.3%) | 1.0 n/a | 3 (0.9%) | 1.0 n/a |
| **Difficulty with ASE/ACE** |  |  |  |  |
| Easy or Very Easy | 330 (89.9%) | 0.77 (0.73 to 0.82) | 310 (90.9%) | 0.71 (0.66 to 0.76) |
| Hard or Very Hard | 37 (10.1%) | 0.65 (0.49 to 0.80) | 31 (9.1%) | 0.58 (0.41 to 0.75) |
| **Pain with ASE/ACE** |  |  |  |  |
| None | 359 (97.8%) | 0.76 (0.71 to 0.80) | 322 (93.9%) | 0.69 (0.64 to 0.74) |
| A little | 6 (1.6%) | 0.83 (0.54 to 1.00) | 17 (5.0%) | 0.76 (0.56 to 0.97) |
| A lot | 1 (0.3%) | 1.0 n/a | 1 (0.3%) | 1.0 n/a |
| I don’t know | 1 (0.3%) | 1.0 n/a | 3 (0.9%) | 1.0 n/a |
| **Ever checked anus for disease** |  |  |  |  |
| No or I don’t know | 196 (53.1%) | 0.74 (0.68 to 0.80) | 189 (54.9%) | 0.69 (0.62 to 0.75) |
| Yes | 173 (46.9%) | 0.79 (0.73 to 0.85) | 155 (45.1%) | 0.72 (0.65 to 0.79) |
| **Worry about getting anal cancer** |  |  |  |  |
| None | 219 (59.7%) | 0.76 (0.70 to 0.81) | 202 (58.9%) | 0.72 (0.66 to 0.78) |
| A little | 101 (27.5%) | 0.75 (0.67 to 0.84) | 103 (30.0%) | 0.65 (0.56 to 0.74) |
| Some | 35 (9.5%) | 0.89 (0.78 to 0.99) | 33 (9.6%) | 0.76 (0.61 to 0.90) |
| Quite a lot | 12 (3.3%) | 0.50 (0.22 to 0.78) | 5 (1.5%) | 0.60 (0.22 to 1.00) |
| **Plans to do ASE/ACE in the future** |  |  |  |  |
| Strongly agree | 257 (70.2%) | 0.78 (0.73 to 0.83) | 251 (73.2%) | 0.70 (0.64 to 0.76) |
| Agree | 99 (27.1%) | 0.71 (0.62 to 0.80) | 85 (24.8%) | 0.71 (0.61 to 0.80) |
| Disagree | 1 (0.3%) | 1.0 n/a | 2 (0.6%) | 1.0 n/a |
| Strongly Disagree | 2 (0.6%) | 0.0 n/a | 0 | 0.0 n/a |
| I don’t know | 7 (1.9%) | 1.0 n/a | 5 (1.5%) | 0.40 (0.00 to 0.83) |
| **Would see a doctor for a persistent anal problem** |  |  |  |  |
| Strongly agree | 245 (66.6%) | 0.76 (0.71 to 0.82) | 236 (68.8%) | 0.70 (0.65 to 0.76) |
| Agree | 96 (26.1%) | 0.82 (0.75 to 0.90) | 87 (25.4%) | 0.69 (0.59 to 0.79) |
| Disagree | 5 (1.4%) | 0.60 (0.17 to 1.00) | 7 (2.0%) | 0.71 (0.38 to 1.00) |
| Strongly Disagree | 3 (0.8%) | 0.33 (0.00 to 0.87) | 0 | 0.0 n/a |
| I don’t know | 19 (5.2%) | 0.53 (0.30 to 0.75) | 13 (3.8%) | 0.69 (0.44 to 0.94) |
| **Preference for ASE/ACE or doctor-provided exam^⸹^** |  |  |  |  |
| ASE or ACE | 121 (33.2%) | 0.80 (0.73 to 0.87) | 123 (36.1%) | 0.70 (0.62 to 0.78) |
| Doctor-provided exam | 243 (66.8%) | 0.74 (0.68 to 0.79) | 218 (63.9%) | 0.71 (0.65 to 0.77) |
| Data are n (%) or median (interquartile range). † Other includes Native American or Alaskan Native, Hawaiian or Pacific Islander, other, and I don’t know. ^*^ Conditions were arthritis, carpal tunnel syndrome, cerebral palsy, diabetes, fibromyalgia, chronic lower back pain, motor neuron diseases, multiple sclerosis, obesity, spina bifida, spinal cord injury, stroke, and other (neuropathy, lower back nerve compression, tremors in hand, McArdle disease, autism, scoliosis, knee pain, scapular dyskinesis, transverse myelitis, osteoporosis, thoracic outlet syndrome, cervicalgia, causalgia and herniated disc). Missing: waist circumference n=6; gender identity n=1; race/ethnicity n=4; sexual orientation n=1; HIV status, self-report n=11; dexterity-related medical condition n=22; recruitment source n=3; preferred anal sex position n=21; difficulty with ASE/ACE n=6; pain with ASE/ACE n=4; ever checked anus for disease n=1; worry about getting anal cancer n=4; Plans to do ASE/ACE in the future n=5; Would see a doctor for a persistent anal problem n=3; Preference for ASE/ACE or doctor-provided exam n=9. Abbreviations: ASE/ACE, anal self-examination, or anal companion examination; n/a, not applicable. | | | | |
| ***Supplemental* *Table 2.* Characteristics of individuals conducting lay anal examinations and concordance with clinician examinations by city in Chicago, Illinois and Houston, Texas, 2020-2022** | | | | |

| **Characteristic** | **PR (95% CI)** | **aPR* (95% CI)** |
| --- | --- | --- |
| **Age**, years |  |  |
| 25-34 | 1.0 | 1.0 |
| 35-44 | 0.91 (0.81-1.03) | 0.92 (0.81-1.04) |
| 45-54 | 0.92 (0.81-1.04 | 0.94 (0.83-1.06) |
| 55-81 | **0.83 (0.73-0.95)** | **0.85 (0.74-0.97)** |
| **Waist**, cm |  |  |
| ≤ 102 | 1.0 |  |
| > 102 | 0.92 (0.83-1.02) | - |
| **Sexual orientation** |  |  |
| Gay | 1.0 |  |
| Bisexual | **1.15 (1.01-1.31)** | - |
| Queer | 1.11 (0.93-1.33) | - |
| Heterosexual, lesbian, don’t know or other | **1.26 (1.02-1.56)** | - |
| **HIV status**, self-report |  |  |
| Negative | 1.0 | - |
| Positive | 0.92 (0.83-1.02) | - |
| **Difficulty with ASE/ACE** |  |  |
| Hard or Very Hard | 1.0 |  |
| Easy or Very Easy | 1.20 (0.99-1.46) | - |
| **Trainer type** |  |  |
| Clinician | 1.0 | 1.0 |
| Non-clinician | **0.84 (0.76-0.93)** | **0.86 (0.77-0.97)** |
| **Clinician type** |  |  |
| Medical doctor | 1.0 |  |
| Advanced practice provider | **0.89 (0.81-0.98)** | **-** |
| **Worried about getting anal cancer** |  |  |
| None | 1.0 | 1.0 |
| A little | 0.95 (0.85-1.06) | 0.97 (0.86-1.08) |
| Some | **1.13 (1.00-1.28)** | **1.14 (1.01-1.29)** |
| Quite a lot | 0.63 (0.37-1.09) | 0.63 (0.37-1.06) |
| Note. Confidence intervals in bold do not include unity. ^*^ Variables remaining in model are adjusted for each other and city. Abbreviations: PR, prevalence ratio; aPR, adjusted prevalence ratio; anal self-examination, or anal companion examination. | | |
| ***Supplemental* *Table 3.* Factors associated with concordance between anal self-examinations and clinician examinations in Chicago, Illinois and Houston, Texas, 2020-2022, bivariate and multivariable analyses** | | |

Assessed for eligibility (n=2,637)

- Declined consent (n=103)

- Non-responsive (n=569)

Consented (n=954)

Completed lay exam (n=714)

- Did not attend first visit (n=236)

- Excluded from analysis (n=4)

- Inadequate DARE (n=1)

- Did not complete exam (n=3)

Not eligible (n=1,011)

Eligible (n=1,626)

***Supplemental Figure.* Study flow.**
